# Correlation of the commercial anti-SARS-CoV-2 receptor binding domain antibody test with the chemiluminescent reduction neutralizing test and possible detection of antibodies to emerging variants

**DOI:** 10.1101/2021.05.25.21257828

**Authors:** Yoshitomo Morinaga, Hideki Tani, Yasushi Terasaki, Satoshi Nomura, Hitoshi Kawasuji, Takahisa Shimada, Emiko Igarashi, Yumiko Saga, Yoshihiro Yoshida, Rei Yasukochi, Makito Kaneda, Yushi Murai, Akitoshi Ueno, Yuki Miyajima, Yasutaka Fukui, Kentaro Nagaoka, Chikako Ono, Yoshiharu Matsuura, Takashi Fujimura, Yoichi Ishida, Kazunori Oishi, Yoshihiro Yamamoto

## Abstract

**Background:** Serological tests are beneficial for recognizing the immune response against SARS-CoV-2. To identify protective immunity, optimization of the chemiluminescent reduction neutralizing test (CRNT), using pseudotyped SARS-CoV-2, is critical. Whether commercial antibody tests are comparably accurate is unknown.

**Methods:** Serum samples collected before variants were locally found were obtained from confirmed COVID-19 patients (n = 74), confirmed non-COVID-19 individuals (n = 179), and unscreened individuals (suspected healthy individuals, n = 229). The convalescent phase was defined as the period after day 10 from disease onset. The CRNT against pseudotyped viruses displaying the wild-type spike protein and a commercially available anti-receptor binding domain (RBD) antibody test were assayed. The CRNT was also assayed, using South African (SA) and United Kingdom (UK)-derived variants.

**Results:** The CRNT (cut off value, 50% inhibition) and the anti-RBD antibody test (cut off value, 0.8 U/mL) concurred regarding symptomatic COVID-19 patients in the convalescent phase and clearly differentiated between patients and suspected healthy individuals (sensitivity; 95.8% and 100%, specificity; 99.1% and 100%, respectively). Anti-RBD antibody test results correlated with neutralizing titer (r = 0.47, 95% CI 0.20-0.68). Compared with the wild-type, CRNT reduction was observed for the SA and UK-derived variants. Of the samples with ≥100 U/mL by the anti-RBD antibody test, 77.8% and 88.9% showed ≥50% neutralization against the UK and the SA variants, respectively.

**Conclusion:** The CRNT and commercial anti-RBD antibody test effectively classified convalescent COVID-19 patients. The strong positive results using the commercial antibody test can reflect neutralizing activity against emerging variants.

## Introduction

Understanding the status of immunity acquisition against severe acute respiratory syndrome coronavirus 2 (SARS-CoV-2) will help us overcome the difficulties related to the coronavirus disease (COVID-19) pandemic. Serological tests can provide information on the immune status after viral exposure and vaccination. The virus neutralizing test is a method for directly determining the functional ability of the immune response; however, it is not suitable as a routine test in clinical laboratories due to its complexity and the risks associated with using live viruses.

We previously established the chemiluminescence reduction neutralization test (CRNT) for the evaluation of immunity to SARS-CoV-2, using pseudotyped virus (1). The CRNT can assess the inhibitory effect of serum samples on viral attachment and entry into the target cells. As observed in our previous reports (1, 2), the inhibition of the infectivity of sera from symptomatic COVID-19 patients gradually increased during the follow-up period, suggesting that the CRNT reflects the status of immunity acquisition. Meanwhile, commercial antibody tests that do not specify functional antibodies are becoming available. Some tests can detect antibodies specific to the receptor-binding domain (RBD) of the spike protein on SARS-CoV-2 that binds to the ACE2 receptor expressed on the targeted cells (3-5). Therefore, any correlation between commercial test results and protective function against SARS-CoV-2 is of epidemiological and clinical interest.

Several SARS-CoV-2 variants have been identified (6). The B.1.351 variant, which was originally identified in South Africa, is characterized by amino acid mutations such as K417N, E484K, and N501Y in the RBD of the spike protein (7). These mutations can be related to the neutralizing function of the immune response of previous COVID-19 as well as viral binding because of structural changes in the contact sites to the ACE2 receptor (8). The B.1.1.7 variant that emerged in the United Kingdom also has the mutation N501Y (9) which has been shown to increase the affinity to the ACE2 receptor (10). It has been suggested that N501Y and the other mutations are not related to the reduction of neutralization (11, 12); however, potentially reduced neutralization has also been implied (13, 14). Thus, elucidating whether antibodies present in the sera from COVID-19 patients have neutralizing activity against emerging SARS-CoV-2 variants remains paramount.

The optimization of immune response tests may help the accurate evaluation of infection-induced antibodies as well as the efficacy of vaccination. The performance of the CRNT to recognize individuals who have likely acquired immunity was evaluated. In addition, we investigated whether the neutralizing effects were reasonably predictable by a commercially available antibody test.

## Materials and Methods

### Collection of specimens

Serum samples were collected from COVID-19 patients, uninfected close contacts, and suspected healthy individuals at the Toyama University Hospital and Toyama City Hospital. The sera were frozen at −80 °C until serological assays were performed. All samples were collected before SARS-CoV-2 variants were found locally.

The diagnosis of COVID-19 was confirmed by the positive results of nasopharyngeal swab samples on the SARS-CoV-2 quantitative reverse transcriptase–polymerase chain reaction (RT-qPCR) testing (confirmed COVID-19 patients). The uninfected close contacts, including health care workers, were those who were tested at least once as negative on the SARS-CoV-2 RT-qPCR (confirmed non-COVID-19 individuals). The suspected healthy individuals (unscreened individuals) were health care workers at the Toyama University Hospital, who were not tested via SARS-CoV-2 RT-qPCR, as they were not considered at risk and did not present with symptoms nor reported close contacts.

Basic clinical characteristics were obtained from medical records or questionnaires from all participants who had undergone SARS-CoV-2 RT-qPCR. The obtained characteristics included symptoms (fever, cough, sputum, sore throat, nasal discharge, loss of taste, loss of smell, dyspnea, and others), underlying diseases (malignant diseases, diabetes, immunosuppression, renal failure, liver failure, and systemic lupus erythematosus), and medication (corticosteroids excluding ointment, immunosuppressants, anti-tumor drugs, anti-rheumatoid drugs, and radiological therapy).

For suspected healthy individuals, medical information on symptoms, underlying diseases, and medications was not collected. The serum samples from these individuals were originally collected in July and August 2020 for the screening of subclinical SARS-CoV-2 infections among staff by the infection control team because at least 3 months had passed since the first case of COVID-19 was hospitalized.

### Virological investigation

SARS-CoV-2 RT-qPCR was performed at officially approved laboratories, including the University of Toyama, Toyama Institute of Health, and external private laboratories. The methods of SARS-CoV-2 RT-qPCR were dependent on each laboratory. Cases of RNAemia were screened (15), and the results were used as the demographic background. When the remaining respiratory specimen were available, co-infected microorganisms were screened by FilmArray Respiratory Panel 2.1 (bioMérieux Japan, Tokyo, Japan), according to manufacturer’s instructions.

### Generation of pseudotyped viruses

Pseudotyped vesicular stomatitis virus (VSV) bearing SARS-CoV-2 S protein was generated as previously described (1). The expression plasmid for the truncated S protein of SARS-CoV-2, pCAG-SARS-CoV-2 S (Wuhan), were kindly provided from Dr. Shuetsu Fukushi, National Institute of Infectious Diseases, Japan. The expression plasmids for the truncated mutant S protein of SARS-CoV-2, pCAGG-pm3-SARS2-Shu-d19-B1.1.7 (UK-derived variant) and pCAGG-pm3-SARS2-Shu-d19-B1.351 (South Africa-derived variant), were constructed by PCR-based site-directed mutagenesis using the cDNA as a template, which obtained by chemical synthesis with optimization for the humanized codon (Thermo Fisher Scientific, MA). The S cDNA of SARS-CoV-2 was cloned into the pCAGGS-pm3 expression vector. Briefly, 293 T cells were transfected with above the expression vectors. After 24 h of incubation, the transfected cells were infected with G-complemented (*G) VSVΔG/Luc (*G-VSVΔG/Luc) at a multiplicity of infection of 0.5. The virus was adsorbed and extensively washed four times with Dulbecco’s modified Eagle’s medium (DMEM) supplemented with 10% fetal bovine serum (FBS). After 24 h of incubation, the culture supernatants containing pseudotyped VSVs were centrifuged to remove cell debris and stored at −80 °C until further use.

### Serological tests

The neutralizing effects of samples against pseudotyped viruses were examined in 96-well microplates (Thermo Fisher Scientific, MA) by the CRNT, as previously described (1). In this study, we utilized VeroE6/TMPRSS2 cells, which were highly susceptible for SARS-CoV-2 infection. VeroE6/TMPRSS2 cells (JCRB1819) was purchased from Japanese Collection of Research Bioresources (JCRB) Cell Bank (Osaka, Japan). Briefly, serum-containing DMEM (Nacalai Tesque, Inc., Kyoto, Japan) with 10% heat-inactivated FBS was incubated with pseudotyped SARS-CoV-2 for 1 h. After incubation, Vero E6 TMPRSS2 cells were treated with DMEM-containing serum and pseudotyped virus. The infectivity of the pseudotyped viruses was determined by measuring the luciferase activity after 24 h of incubation at 37 °C. For the high-throughput assay (htCRNT), the CRNT was modified, using 384-well microplates (Corning, NY).

For the commercial assay, the serum samples were tested at an external laboratory company, using Elecsys Anti-SARS-CoV-2 S immunoassay (Roche Diagnostics GmbH, Basel, Switzerland) to quantitatively measure the antibody levels to the SARS-CoV-2 RBD. The manufacturer’s cut-off value (COV) was 0.8 U/mL and the minimum value was expressed as <0.4 U/mL.

### Statistical analysis

Statistical analysis was performed using the Kruskal-Wallis test with Dunn’s test for multiple comparisons among three groups or more. Correlations between test findings were expressed using Pearson’s correlation coefficients. Positive conversion was analyzed by the Kaplan-Meier method, using the Gehan-Breslow-Wilcoxon test. Data were analyzed, using GraphPad Prism version 8.4.3 (GraphPad Software, CA). Statistical significance between different groups is presented in the figure legend.

### Ethics approval

This study was performed in accordance with the Declaration of Helsinki and was approved by the ethical review board of the University of Toyama (approval No.: R2019167 and R2020097). Written informed consent was obtained from all participants.

## Results

To investigate the relationship between the clinical findings and seroconversion, 482 serum samples, excluding three samples with low volume remaining, were evaluated. These samples were collected from confirmed COVID-19 patients (n = 74), confirmed non-COVID-19 individuals (n = 179), and unscreened individuals (suspected healthy individuals) (n = 229) (**Table 1**). Because in our previous study, moderate and severe patients with COVID-19 showed > 50% inhibition (IC_50_) in the CRNT after day 10 from disease onset (2), the period after day 10 from disease onset was considered as the convalescent phase. Neutralization activity against pseudotyped viruses and anti-RBD antibody levels were evaluated by the CRNT (**Figure 1A**) and quantified by the commercially available test (**Figure 1B**). Because asymptomatic individuals can have a weak immune response to SARS-CoV-2 infection (16), the diagnostic performance of both tests was evaluated by comparing the symptomatic confirmed COVID-19 patients in the convalescent phase (n = 24) and the unscreened individuals (**Figure 1C**). Both tests clearly classified these two groups (the best COVs, CRNT; 50.5, anti-RBD antibody test; 0.62). Thus, in the following analysis, IC_50_ for the CRNT and 0.8 U/mL for the anti-RBD antibody test (manufacturer’s COV) were used as the COVs for predicting serological positive conversion to SARS-CoV-2.

**Table 1.** Demographic and clinical characteristics of the study participants.

| Profile | Confirmed<br>COVID-19, n=74 | Confirmed<br>non-COVID-19,<br>n=179 | Unscreened,<br>n=229 |
| --- | --- | --- | --- |
| Sex, male, n (%) | 32 (43.2) | 60 (33.5) | 101 (44.1) |
| Age, years, n (%) |  |  |  |
| ≤19 | 4 (5.4) | 1 (0.6) | 0 (0.0) |
| 20-29 | 14 (18.9) | 42 (23.5) | 51 (22.3) |
| 30-39 | 11 (14.9) | 51 (28.5) | 71 (31.0) |
| 40-49 | 6 (8.1) | 39 (21.8) | 53 (23.1) |
| 50-59 | 10 (13.5) | 25 (14.0) | 35 (15.3) |
| 60-69 | 9 (12.2) | 11 (6.1) | 19 (8.3) |
| ≥70 | 20 (27.0) | 10 (5.6) | 0 (0.0) |
| Symptom, n (%) |  |  |  |
| Symptomatic | 66 (89.2) | 56 (31.3) | NA |
| Fever | 50 (67.6) | 22 (12.3) |  |
| Cough | 43 (58.1) | 22 (12.3) |  |
| Sputum | 18 (24.3) | 17 (9.5) |  |
| Sore throat | 15 (20.3) | 19 (10.6) |  |
| Nasal discharge | 7 (9.5) | 13 (7.3) |  |
| Loss of taste | 22 (29.7) | 2 (1.1) |  |
| Loss of smell | 25 (33.8) | 3 (1.7) |  |
| Dyspnea | 21 (28.4) | 11 (6.1) |  |
| Others | 19 (25.7) | 12 (6.7) |  |
| Asymptomatic | 8 (10.8) | 123 (68.7) | NA |
| Phase of blood sampling, n (%) |  |  |  |
| Acute (< 9 days from onset or a close contact episode), n (%) | 48 (64.9) | 5 (2.8) | NA |
| Convalescent (≥10 days from onset or a close contact episode), n (%) | 26 (35.1) | 174 (97.2) | NA |
| Underlying diseases, n (%) |  |  |  |
| Yes | 14 (18.9) | 10 (5.6) | NA |
| Malignant diseases | 2 (2.7) | 4 (2.2) |  |
| Diabetes | 8 (10.8) | 7 (3.9) |  |
| Immunosuppression | 3 (4.1) | 1 (0.6) |  |
| Renal failure | 5 (6.8) | 2 (1.1) |  |
| Liver failure | 0 (0.0) | 1 (0.6) |  |
| Systemic lupus erythematosus | 0 (0.0) | 0 (0.0) |  |
| No | 60 (81.1) | 169 (94.4) | NA |
| Medication, n (%) |  |  |  |
| Yes | 5 (6.8) | 7 (3.9) | NA |
| Corticosteroids (excluding ointment) | 3 (4.1) | 6 (3.3) |  |
| Immunosuppressants | 1 (1.4) | 1 (0.6) |  |
| Anti-tumor drugs | 0 (0.0) | 0 (0.0) |  |
| Anti-rheumatoid drugs | 1 (1.4) | 1 (0.6) |  |
| Radiological therapy | 0 (0.0) | 1 (0.6) |  |
| No | 69 (93.2) | 172 (96.1) | NA |

NA, not applicable

**Figure 1.**
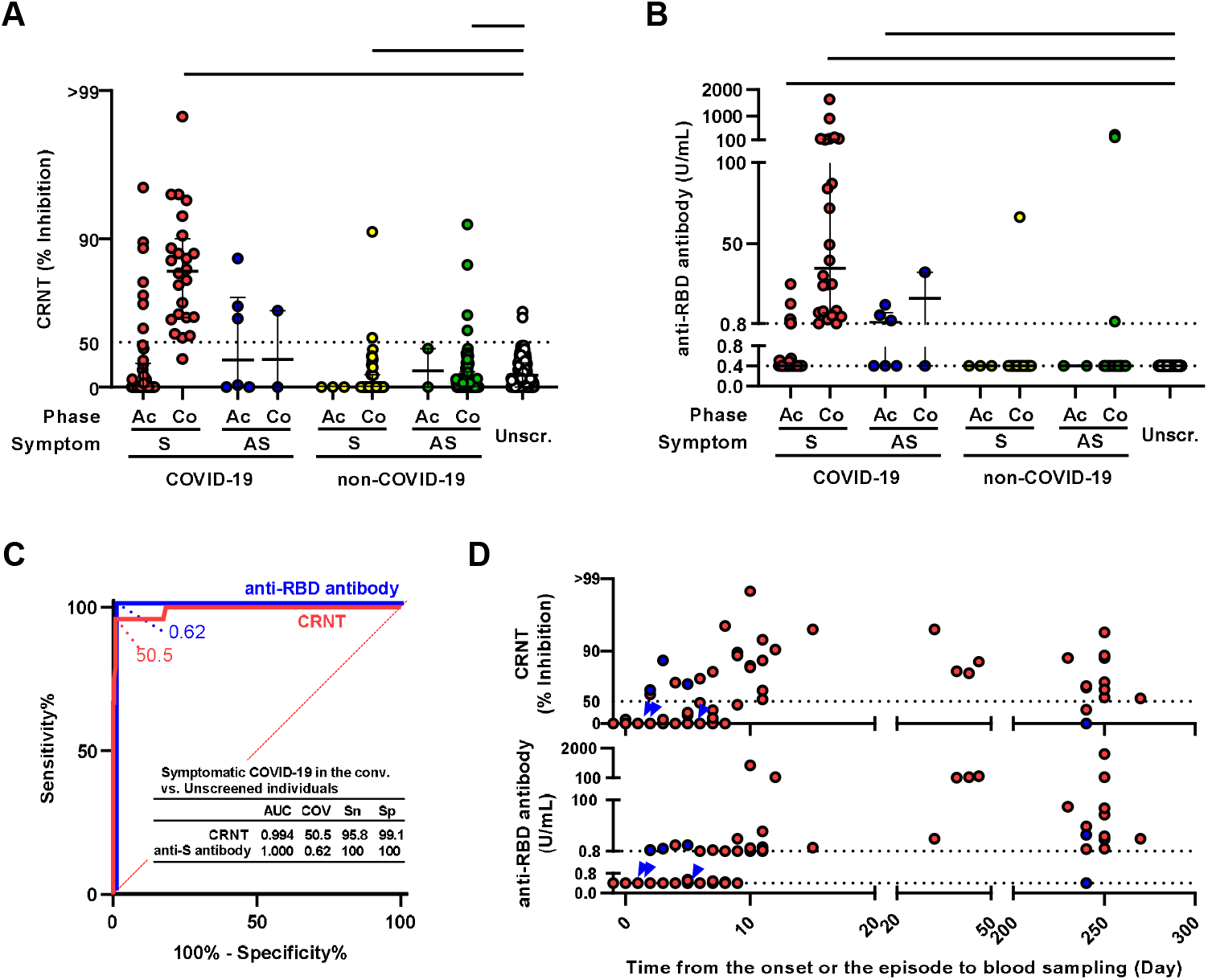
Neutralization and anti-RBD antibody levels. **(A)** Neutralization levels against pseudotyped viruses measured by CRNT (serum dilution, x 100). **(B)** Anti-RBD antibody levels measured by commercially available test. **(C)** ROC curves to classify the symptomatic confirmed COVID-19 patients in the convalescent phase and the unscreened individuals. **(D)** Relationship between results of each test and time from symptom onset or a close contact episode to blood sampling in COVID-19 patients. Symptomatic and asymptomatic individuals are presented in red and blue (blue arrowhead for overlapping cases), respectively. *** indicates p < 0.001 by unpaired Kruskal-Wallis test and Dunn’s multiple comparison with setting the unscreened group as a control. Ac, acute phase; Co, convalescent phase; S, symptomatic; AS, asymptomatic; Unscr., unscreened; AUC, area under the curve; COV, cut-off value; Sn, sensitivity; Sp, specificity.

For the CRNT (**Figure 1A** and **Supplemental Table 1**), the percentage of positivity in the symptomatic confirmed COVID-19 patients was 16.7% (7/42) in the acute phase and 95.8% (23/24) in the convalescent phase. The CRNT values of the symptomatic confirmed COVID-19 patients (median 83.5; IQR 64.1-90.0) were significantly higher than those of the unscreened individuals (% positivity: 0.9% [2/229], median 17.0; IQR 0.0-31.2) (p < 0.001). Conversely, symptomatic and asymptomatic confirmed non-COVID-19 individuals in the convalescent phase showed significantly decreased CRNT values (p < 0.001).

For anti-RBD antibody levels (**Figure 1B** and **Supplemental Table 1**), in the symptomatic confirmed COVID-19 patients, 14.3% (6/42) in the acute phase and 100.0% (24/24) in the convalescent phase tested positive. In contrast, 0.0% (0/229) of the unscreened individuals were positive. Compared with the unscreened group, symptomatic confirmed COVID-19 patients in the acute phase, those in the convalescent phase, and asymptomatic confirmed COVID-19 patients in the acute phase showed significant increases in their serum anti-RBD antibody levels (median 0.40; IQR 0.40-0.40, median 0.40; IQR 0.40-0.46, median 35.0; IQR 7.63-137.0, and median 1.59; IQR 0.40-7.55, respectively, p < 0.001).

Among the confirmed COVID-19 patients, the positivity of both tests was time-dependent (**Figure 1D**). Positive conversion was observed 2 days after the onset or the episode of close contact, and cases in the convalescent phase were positive for both tests, excluding two cases sampled after 240 days. While the confirmed COVID-19 group included sub-populations with RNAemia analyzed, it was independent of positive conversion of the neutralization or anti-RBD antibody during the acute phase (**Supplemental Figure 1**).

### Relationship of the anti-RBD antibody test and neutralizing response

To evaluate the indirect functional meaning of the anti-RBD antibody test, these values were compared with those of the CRNT. The concordance of the CRNT and the anti-RBD antibody test was 98.9% (477/482; double positive: n = 38; double negative: n = 439) (**Figure 2A**). Of five samples with discordance, four were slightly positive for CRNT (53.5-69.0) but negative for anti-RBD antibody test (< 0.40 U/mL). For the other discordance, of which the value of anti-RBD antibody test was slightly elevated (5.06 U/mL), CRNT was partly inhibited (CRNT 35.5) but judged as negative. Discordance was not related to the underlying diseases or medications.

**Figure 2.**
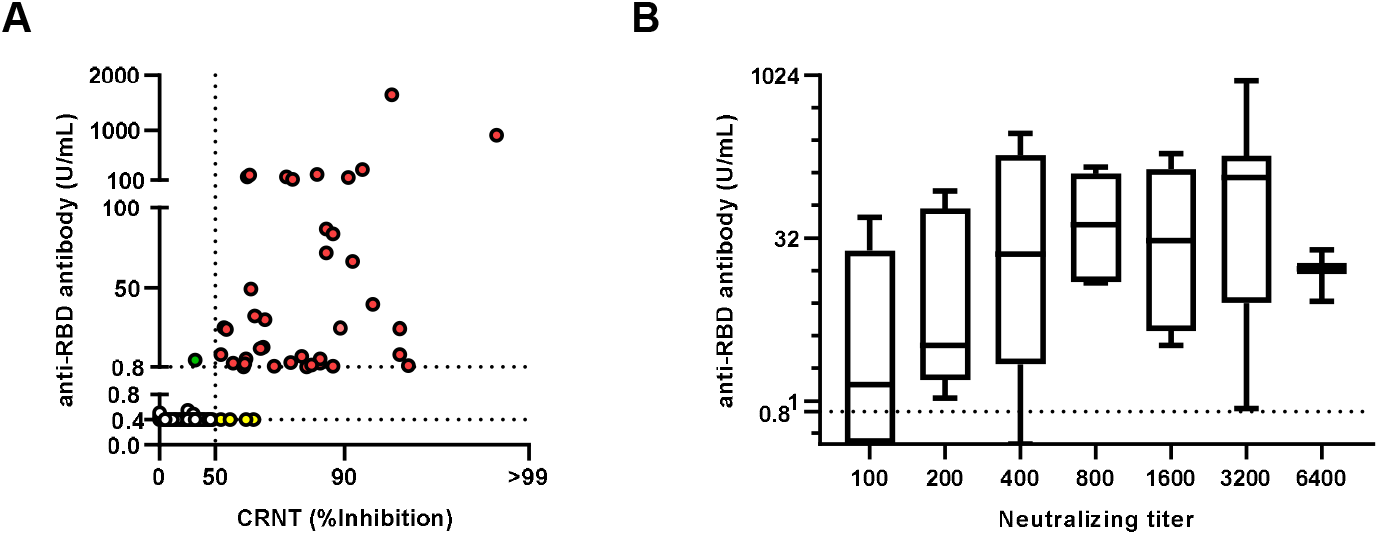
Relationship between CRNT and anti-RBD antibody test. **(A)** Comparison of neutralization levels and anti-RBD antibody results. Concordant samples are red (positive for both tests) or white (negative for both tests). Discordant samples are green (positive for anti-S antibody) or yellow (positive for CRNT). Dotted line of CRNT indicates 50% of infectivity (IC_50_). **(B)** Values of anti-RBD antibody test and neutralizing activity. Sera positive for CRNT (serum dilution, x100) were serially diluted up to x6400 and the dilution with >IC_50_ was defined as a neutralizing titer. Box indicate median and interquartile. Error bars indicate minimum to maximum.

Next, to evaluate whether the values measured by the anti-RBD antibody test indirectly correlated with neutralizing activity, these values were compared with CRNT using the diluted sera (**Figure 2B**). The values positively correlated with neutralizing titer (r = 0.47, 95% confidence interval 0.20-0.68); the sera with higher values tended to be positive for CRNT (> 50% inhibition) despite being highly diluted.

### Performance of high throughput assay

To increase the number of simultaneous processing, the performance of htCRNT, which is a high-throughput neutralizing assay on a 384-well plate, was also evaluated using 100-fold-diluted sera. The results of htCRNT correlated with those of CRNT (r = 0.83, 95% confidence interval 0.80-0.86), and concordance was 98.8% (476/482). Six discordant samples were negative for htCRNT but positive for CRNT (**Supplemental Figure 2A** and **Supplemental Table 2**). Significant inhibition was observed in symptomatic confirmed COVID-19 in the convalescent phase (median 83.5; IQR 67.7-91.0) compared to the unscreened individuals (median 0.0; IQR 0.0-11.0, p < 0.001, **Supplemental Figure 2B**). The cut-off value was set as IC_50_ based on the ROC analysis (**Supplemental Figure 2C**), and the concordance of the two tests was 99.4% (479/482; double positive: n = 36; double negative: n = 443, **Supplemental Figure 2D**). Three discordant samples were positive for the anti-RBD antibody test but negative for the htCRNT (htCRNT 28-41). Three cases in the convalescent phase were negative for the htCRNT (**Supplemental Figure 2E**).

### Evaluation of cross reaction to pseudotyped SARS-CoV-2 variants

Finally, to investigate the neutralizing activity against SARS-CoV-2 variants, CRNT-positive samples were assayed using pseudotyped SA and UK-derived variants. Compared to the wild-type pseudotyped virus (Wuhan), the CRNT values against the UK and SA-derived variants were significantly decreased (median 80.8; IQR 66.4-88.8 for WT, vs. median 53.4; IQR 36.7-67.4; median 43.1; IQR 14.2-60.2, respectively) (**Figure 3A**). The percentages of serum samples above CRNT 50.0 were 77.8% (7/9) for UK-derived variant and 88.9% (8/9) for SA-derived variant among samples with ≥ 100 U/mL by anti-RBD antibody test, while those were 53.6% (15/28) for UK-derived variant and 32.1% (9/28) for SA-derived variant among samples with 0.8-<100 U/mL by the anti-RBD antibody test (**Figure 3B**).

**Figure 3.**
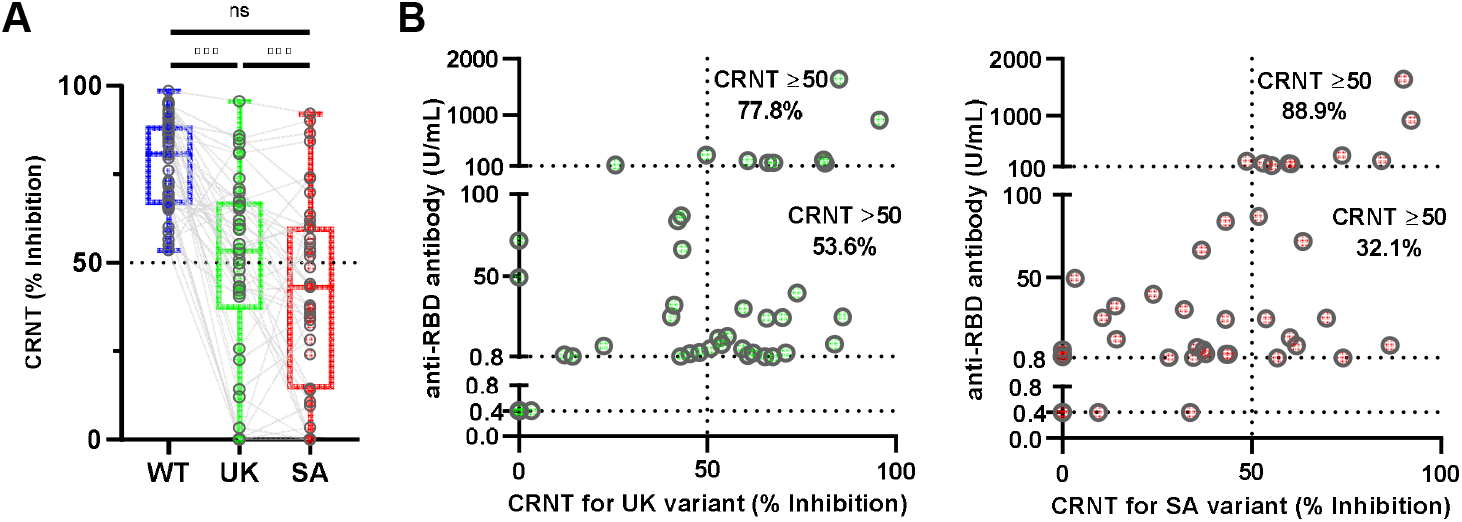
Neutralizing activities against SARS-CoV-2 variants in Wuhan-CRNT-positive sera. **(A)** Neutralizing sensitivity of SARS-CoV-2 pseudotyped variants. Neutralization of wild-type (WT) spike protein (Wuhan) CRNT-positive sera (serum dilution, x100) was assessed, using pseudotyped viruses displaying the mutant spike proteins (UK and SA-derived variants). Box indicate median and interquartile. Error bars indicate minimum to maximum. **(B)** Relationship between values of anti-RBD-antibody test and neutralization against UK and SA-derived variants (serum dilution, x100).

## Discussion

The commercial antibody test used in the present study showed excellent correlation with the CRNT, suggesting that it is helpful to speculate on the protective antibody response. Because it can detect antibodies specific to the RBD, the values may reflect the number of antibodies with protective functions against SARS-CoV-2 (17, 18). Similarly, assays detecting antibodies against the RBD protein have shown better correlation with neutralizing activity than those against the N protein (4); however, the correlation depends on the manufacturer (3, 4). Our findings suggest that CRNT and anti-RBD antibody tests are suitable for detecting SARS-CoV-2 infection. Seropositive individuals in the non-COVID-19 and unscreened groups might acquire an immune response without noticing SARS-CoV-2 infection or cross-react with other antigens. However, the serological response against SARS-CoV-2 is not yet fully understood. Cross-reactivity against SARS-CoV-2 spike protein may exist (19, 20) whereas normal intravenous immunoglobulin collected before the COVID-19 pandemic has no direct effect against SARS-CoV-2 (21).

Meanwhile, it remains unclear what value may indicate reasonable protection from the future risk of SARS-CoV-2 exposure. It is important to know whether seropositivity can protect against secondary exposure to SARS-CoV-2 (22). To confirm this effect, longitudinal observations are required. However, the values of the anti-RBD antibody test and CRNT showed a positive correlation, which is consistent with previous reports (3, 23). While not all antibodies against the RBD are neutralizing (24), the quantitative value of the anti-RBD antibody test may partly reflect the prophylactic function.

The long-term sustainability of effective immunity after recovery from COVID-19 remains controversial. While early reports indicated sustained humoral immunity in the sera from previously infected patients (18, 25), SARS-CoV-2 reinfection has been reported despite the development of humoral immune response after primary infection (26, 27). Although neutralizing antibodies may disappear after 6 months or longer after the onset (4), in the present study, most of the convalescent sera from COVID-19 patients showed inhibition of viral infection. Asymptomatic COVID-19 patients may have a weaker immune response than symptomatic patients or show a relative reduction in anti-RBD antibody and neutralizing antibody levels, as previously reported (16). These findings support the evidence of long-term immunity against SARS-CoV-2, in particular, in symptomatic patients; however, continuous precautions, including infection control and prevention and vaccination are required for reliable immunity, as convalescent sera can contain insufficient levels of neutralizing activity (25).

Cross-reactivity to SARS-CoV-2 variants was clinical interest. In the present study, the reduction in neutralization was observed for the SA and UK-derived variants compared with the wild-type. In previous reports, neutralization against the UK variant including UK-derived variant were similar when compared with those to the wild-type (11, 28, 29). In contrast, the reduction in neutralization against the UK variant has been also observed in previous reports (13, 14, 29). Our findings suggest that the neutralization against UK variant cannot be completely similar to wild-type, supporting the systematic report (29). For the SA-derived variant, neutralization was significantly reduced as previously reported (11, 28, 29). However, most of sera reacted at least slightly to the SA pseudotyped virus even though the CRNT values were smaller than those associated with the wild-type pseudotyped virus, suggesting that they had a partial cross-reaction. In addition, cross-reactivity was partly reflected in the values obtained with the commercial anti-RBD antibody test. These findings imply that the total amount of antibodies against SARS-CoV-2 RBD can serve as an indirect indicator of a protective role against these variants.

The microassay, htCRNT, showed a good correlation with the CRNT. Because it requires a smaller volume of serum and virus solution than the standard CRNT, a large number of samples can be simultaneously assayed in a plate. Furthermore, because the neutralizing antibody test is the gold standard for assessing functional immune status against the virus, it can provide evidence for planning the recovery of social activity. There are many issues regarding immune status, such as the antibody response of healthcare workers after vaccination, and the antibody retention ratio in the community. Therefore, a high-throughput option can provide an opportunity to conduct large-scale screening.

There are several limitations to the present study. First, serum samples in the present study were one-time collections. Therefore, the continuous antibody level trend and its relationship with disease severity could not be evaluated. Second, sera from individuals who had no evidence of infection, such as sera before the COVID-19 pandemic, could not be set as a control. Lastly, it is unknown whether cross-reactivity to variants is also observed in other commercial antibody tests. Because antigen-antibody relationships are specific, the test performance should be evaluated individually.

The CRNT and anti-RBD antibody tests efficiently detect convalescent COVID-19 patients. Because most facilities cannot evaluate neutralizing antibodies, the good correlation of the non-functional antibody test with the CRNT may help assess the levels of functional antibodies in COVID-19 patients and vaccinated individuals.

## Data Availability

All available data are in the figures and Tables including supplementary data.

## List of abbreviations

CRNT: Chemiluminescent reduction neutralizing test
RBD: Receptor binding domain
RT-qPCR: Quantitative reverse transcriptase–polymerase chain reaction
VSV: Vesicular stomatitis virus
DMEM: Dulbecco’s modified Eagle’s medium
FBS: Fetal bovine serum
COV: Cut-off value

## Acknowledgments

We thank all the staff at Toyama University Hospital and Toyama City Hospital for their help in collecting specimens. We also thank Yumiko Nakagawa and Yoriko Ito for their secretarial assistance. We would like to thank Mayu Somekawa for the arrangement of the specimens.

## Contribution

Conceptualization YoMo

Methodology YoMo, HT

Validation YoMo, HT

Formal Analysis YoMo

Investigation YoMo, HT, HK, TS, EI, YS, MK, and YuMu (neutralizing assay)

YoYo, RY (viral investigations)

Resources HT, TS, EI, YS (generating pseudotyped viruses)

CO, YoMa (generating plasmids)

YT, SN, HK, MK, YuMu, AU, YuMi, YF, and KN (collecting serum samples)

Data Curation YoMo, HT

Writing – Original Draft Preparation YoMo, HT

Writing – Review and Editing YoMo, HT, YU, AS, YK

Visualization YoMo

Supervision TF, YI, KO, YoYa

Project Administration YoMo, YoYa

Funding YoMo, HT, YoYa

---

**Supplementary Table 1.** Basic statistics of SARS-COV-2 serological tests.

| Profile | Confirmed COVID-19 |  |  |  | Confirmed non-COVID-19 |  |  |  | Unscreened |
| --- | --- | --- | --- | --- | --- | --- | --- | --- | --- |
|  | symptomatic |  | asymptomatic |  | symptomatic |  | asymptomatic |  |  |
|  | acute | convalescent | acute | convalescent | Acute | convalescent | acute | convalescent |  |
| N | 42 | 24 | 6 | 2 | 3 | 53 | 2 | 121 | 229 |
| CRNT (%) |  |  |  |  |  |  |  |  |  |
| Minimum | 0.0 | 35.5 | 0.0 | 0.0 | 0.0 | 0.0 | 0.0 | 0.0 | 0.0 |
| 25% Percentile | 0.0 | 64.1 | 0.0 | 0.0 | 0.0 | 0.0 | 0.0 | 0.0 | 0.0 |
| Median | 0.0 | 83.5 | 34.5 | 34.7 | 0.0 | 0.0 | 22.5 | 0.0 | 17.0 |
| 75% Percentile | 30.6 | 90.0 | 75.2 | 69.5 | 0.0 | 17.5 | 45.0 | 21.2 | 31.2 |
| Maximum | 95.5 | 98.5 | 86.5 | 69.5 | 0.0 | 91.0 | 45.0 | 92.0 | 19.0 |
| htCRNT (%) |  |  |  |  |  |  |  |  |  |
| Minimum | 0.0 | 28.0 | 0.0 | 10.0 | 0.0 | 0.0 | 0.0 | 0.0 | 0.0 |
| 25% Percentile | 0.0 | 67.2 | 0.0 | 10.0 | 0.0 | 0.0 | 0.0 | 0.0 | 0.0 |
| Median | 0.0 | 83.5 | 45.5 | 25.5 | 0.0 | 0.0 | 2.0 | 0.0 | 0.0 |
| 75% Percentile | 15.0 | 91.0 | 86.0 | 31.0 | 0.0 | 0.0 | 4.0 | 0.0 | 11.0 |
| Maximum | 98.0 | 98.0 | 89.0 | 31.0 | 0.0 | 83.0 | 4.0 | 91.0 | 50.0 |
| Anti-RBD antibody (U/mL) |  |  |  |  |  |  |  |  |  |
| Minimum | 0.40 | 0.84 | 0.40 | 0.40 | 0.40 | 0.40 | 0.40 | 0.40 | 0.40 |
| 25% Percentile | 0.40 | 7.63 | 0.40 | 0.40 | 0.40 | 0.40 | 0.40 | 0.40 | 0.40 |
| Median | 0.40 | 35.0 | 1.59 | 16.4 | 0.40 | 0.40 | 0.40 | 0.40 | 0.40 |
| 75% Percentile | 0.46 | 137.0 | 7.55 | 32.4 | 0.40 | 0.40 | 0.40 | 0.40 | 0.40 |
| Maximum | 25.1 | 1640 | 12.3 | 32.4 | 0.40 | 66.4 | 0.40 | 294.0 | 0.40 |

CRNT; chemiluminescent reduction neutralizing test, htCRNT; high-throughput CRNT, RBD; receptor binding domain

**Supplemental Figure 1.**
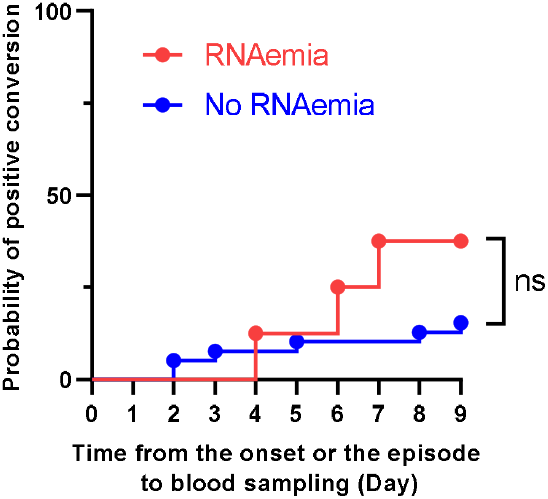
Relationship between seroconversion and RNAemia in confirmed COVID-19 patients in the acute phase. The Kaplan-Meier analysis was performed only for patients who had RNAemia. ns; not significant.

**Supplemental Figure 2.**
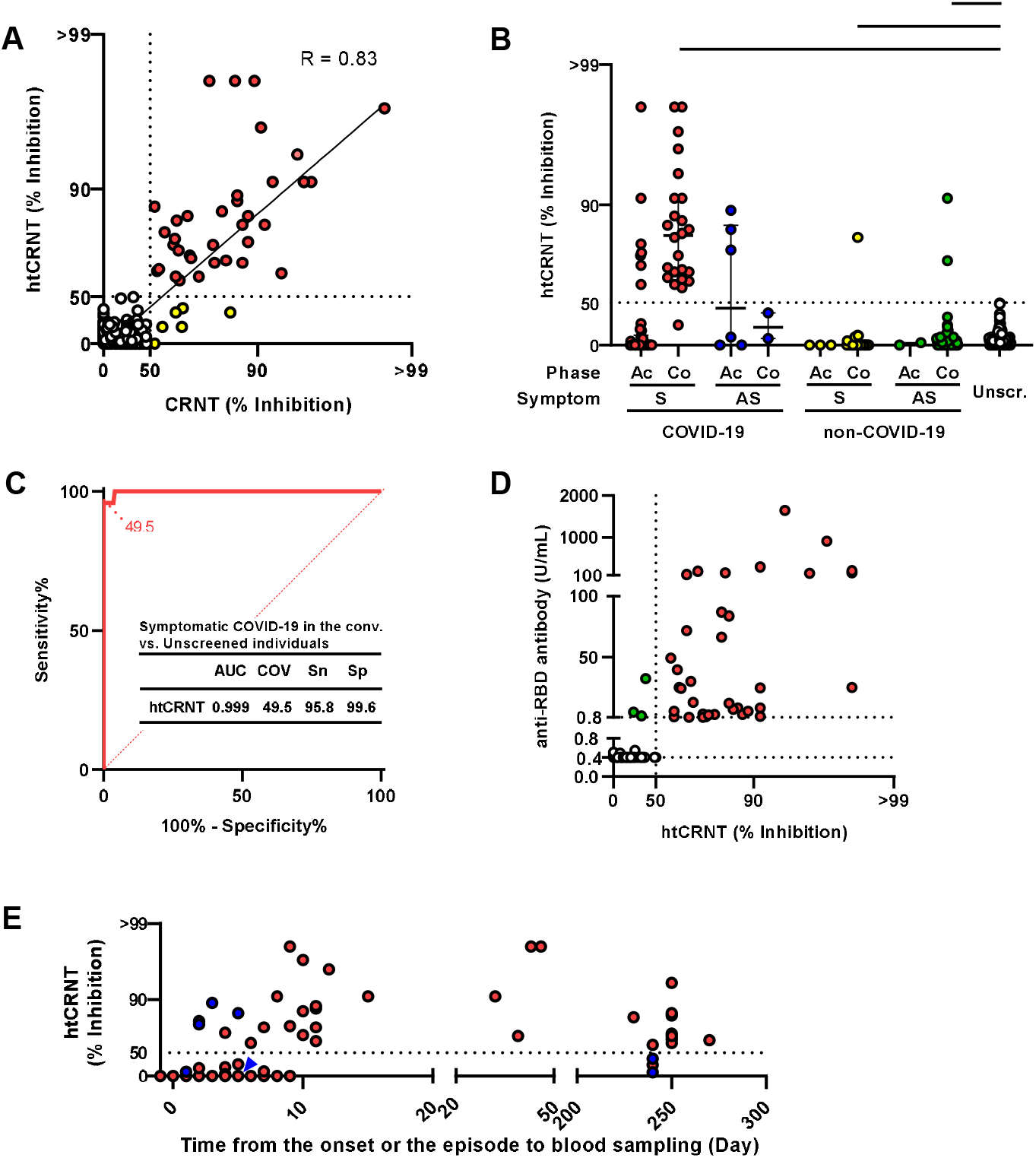
**(A)** Correlation between htCRNT and CRNT. Neutralization was assessed using x100-diluted serum samples. Correlations were expressed using Pearson’s correlation coefficients. **(B)** Neutralization levels against pseudotyped viruses measured, using htCRNT. **(C)** ROC curve of htCRNT for classification of symptomatic confirmed COVID-19 patients in the convalescent phase and unscreened individuals. **(D)** Comparison of htCRNT neutralization levels and anti-S antibody results. Concordant samples were red (positive for both tests) or white (negative for both tests). Discordant samples were green (positive for anti-S antibodies). The dotted line of htCRNT indicates 50% inhibition (IC_50_). **(E)** Relationship between the results of htCRNT and time from symptom onset or a close contact episode to blood sampling in COVID-19 patients. Symptomatic and asymptomatic individuals are presented in red and blue (blue arrowhead for overlapped), respectively. Error bars indicate medians (middle lines) and interquartile range. *** indicates p< 0.001 by unpaired Kruskal-Wallis test and Dunn’s multiple comparison (unscreened group as a control). Ac, acute phase; Co, convalescent phase; S, symptomatic; AS, asymptomatic; Unscr., unscreened; AUC, area under the curve; COV, cut-off value; Sn, sensitivity; Sp, specificity.

